# Multidisciplinary approach to COVID-19 risk communication: A framework and tool for individual and regional risk assessment

**DOI:** 10.1101/2020.07.11.20151464

**Authors:** Rishi Ram Parajuli, Bhogendra Mishra, Amrit Banstola, Bhoj Raj Ghimire, Shobha Poudel, Kusum Sharma, Sameer Mani Dixit, Sunil Kumar Sah, Padam Simkhada, Edwin Van Teijlingen

## Abstract

The COVID-19 pandemic has exceeded over ten million cases globallywith no vaccine available yet. Different approaches are followed to mitigate its impact and reduce its spreading in different countries, but limiting mobility and exposure have been de-facto precaution to reduce transmission. However, a full lockdown cannot be sustained for a prolonged period. Evidence-based, multidisciplinary approach on risk zoning, personal and transmission risk assessment on a near real-time, and risk communication would support the optimized decisions to minimize the impact of coronavirus on our lives. This paper presents a framework to assess the individual and regional risk of COVID-19 along with risk communication tools and mechanisms. Relative risk scores on a scale of 100 represent the integrated risk of influential factors. The personal risk model incorporates: age, exposure history, symptoms, local risk and existing health condition, whereas regional risk is computed through the actual cases of COVID-19, public health risk factors, socioeconomic condition of the region, and immigration statistics. A web application tool (www.covira.info) has been developed, where anyone can assess their risk and find the guided information links primarily for Nepal. This study provides regional risk for Nepal, but the framework is scalable across the world. However, personal risk can be assessed immediately from anywhere.

## Introduction

Coronavirus disease (COVID-19) outbreak originating in China in late 2019 has spread worldwide claiming 1/2 a million lives[1], [2]. The rapid rate of human to human transmission of the virus has threatened the health and livelihood of the entire world[3]. The World Health Organization (WHO) declared it a pandemic on 11^th^ March 2020[1]. Containment and quarantine of the virus contraction have been the major tool to control the spreading in early stage, however, the increasing rate of infections shown the limitations of this approach[4]. Several countries enforced strict lockdown to limit the spread of COVID-19, such as Italy and Spain, or the during the initial phase of the outbreak as in Nepal[5]. WHO issued guidelines for responding to viruses prioritizing actions including maintaining hospital facilities, raising public awareness, and stocking up medical supplies[6].

Public awareness of causative factors of COVID-19, the intensity, risk level, and its consequences could help movivate people to adopt the required public health measures rather than ignoring or over-reacting during this pandemic. A preliminary study from China showed several psychological consequences arising during the COVID-19 pandemic[7]. Appropriate risk perception and communication could greatly help in the management of fear and knowledge sharing during pandemic[8], [9]. To predict the most likely scenarios, as governments and as individuals, the right tools need to be available.

Institutional and individual behaviour in society greatly affects the spread of infectious diseases[10]. Containment, contact tracing, and testing of infectious people should be prioritiesin the early stages of the pandemic [11]. Several developing countries like Nepal introduced strict lockdown measures during the early phase of the pandemic,, but still failed to control the spreading of the virus. Even after a long 21/2 months of lockdown, new infections are mounting up[12]. Citizen and institutional/governance awareness is always a key component in disaster risk reduction, something which is often lacking in low-income countries[13].

Limiting economic activities to combat the pandemic have multi-dimensional impacts; increasing economic uncertainties and exacerbating the vulnerability of the improvised community[14]. Demand and supply chains all over the world have been disrupted[15]. United Nations (UN) agencies have reported that lockdown in some low-income countries resulted in hunger and poverty [16], [17].There needs to be a balace between conroling COVID-19 and maintaining economic activity including the supply of food and essentials though it is challenging[18].

A personal and spatial risk assessment is required to cope with increasing challenges of the absolute lockdown. It paves the way to open the essential operations in the low-risk area by providing safety guidelines about the precaution measures that could fuel the socio-economy of the local community. Personal and regional risk assessment is a systematic and scientific way, in near-real-time (daily basis) to guide individuals and communities to take the appropriate decision. This is more effective in the countries like Nepal, where the current transmission pattern of COVID-19 is sporadic as early COVID-19 cases required the virus abroad with subsequent local transmission. Regular personalized risk indicator and spatial risk zonation not only provides up-to-date information but also change the behaviour in risk perception which could effectively be used to ease the strict lockdown measures in badly hit countries [19], [20].

In this paper, we propose a framework for the COVID-19 risk assessment by incorporating the COVID-19 cases, exposure, immigration (quarantined data), public health facility, and population density, to access the regional and personal risk. We developed a near real-time COVID-19 Risk Assessment (COVIRA) tool based on the proposed framework. The COVIRA incorporates the virus transmission rate, public health risk, and population vulnerability, socio-economic status of the region, and importance of the region in overall context for essentials production and supply. Additionally, personal risk assessment provides individual vulnerability and risk of COVID-19 infection. Personalized risk communication could support limiting the spread of the virus and ultimately provide a better way to exit the pandemic. As, the personal, and regional risk computed through COVIRA reflect the recent scenario, an appropriate measure can be taken to remain safe from the infection and bring the life of people to normal as soon as possible.

## Result

The COVIRA is a dynamic system for regional and personal COVID-19 risk assessment based on the most recent data available. Results are presented in three segments; personal risk, regional risk assessment, and zonal importance to provide the overall scenario of COVID-19 risk in real-time. Personal risk provides the individual vulnerability towards this pandemic which will notifythe risk of: (i) an individual getting infected, (ii) COVID-19 infection and (iii) being infected through regional risk. Regional risk assessment provides COVID-19 transmission risk in the region, public health risk, socioeconomic risk and the overall risk. One major aspect of managing exit from the pandemic is prioritizing essentialservices by providing the regional importance maps and guidance. This would effectively be updated to guide the community and individual when the government policy changes for the priorities during different stages of a pandemic. All risk factors are normalized on a scale of 100 in the calculation.

### Personal risk assessment

Age and existing health conditions are two major factors contributing to COVID-19 risk. Data analysis from six countries (China, Italy, Spain, Germany, UK [United Kingdom] and USA [United States of America]) provide a fair relationship of these factors in patients’ death. Age is exponentially correlated with risk where underlying health conditions are contributing nearly 93% of overall death in UK and USA. Data suggest that the older people are at high risk from COVID-19. A fraction of deaths to total deaths in each age group and percentage of -non-survivors to total positive cases in corresponding age groups, both indicate an almost similar trend, where death percentage of total positive cases promisingly shows the exponential relationship with age as shown in equation 1. Data correlation between each country is shown in methodology with more details on appendix from both perspectives.

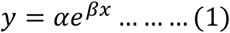

Where, ‘y’ is a normalized risk factor; ‘x’ is the age in years (up to 90 years), normalized by mean 45.11 and standard deviation 27.32; α and β are coefficients having values (95% confidence level) 8.947 (5.955, 11.94) and 1.492 (1.272, 1.712) respectively; R-squared value of this relationship is 0.9538.

Underlying health condition from China[21], [22], the USA[23] and the UK[24] are also analyzed to find the correlation of different underlying health conditions over the age due to COVID-19. Guan et al.[21] reported total cases and severe cases, where Zhou et al.[22] reported total cases and corresponding deaths associated with different underlying health conditions. Garg et al.[23] reported only hospitalization rates of several underlying health conditions in different age groups. The UK data has been published by Docherty et al.[24] reported a total of 16749 patients data with different symptoms and underlying health conditions. Average of relative risk factors of different existing health conditions, comorbidities risk factor (CRF) from available data are provided in Table 1, detailed data can be found in appendix.

**Table 1.** Relative risk of existing health condition to COVID-19.

| Comorbidities | CRF |
| --- | --- |
| Asthma/ Chronic obstructive pulmonary disease | 100 |
| Cardiovascular/Coronary heart disease | 99 |
| Renal disease | 90 |
| Diabetes | 69 |
| Hypertension | 54 |
| Obesity | 54 |
| Cancer | 54 |
| Cerebrovascular disease | 48 |

Following the trend of percentage of deaths with an existing health condition to the different age groups, another function (z) for a coefficient of comorbidity has been derived from UK data[25] as shown in equation 2. Equation 3 provides total relative risk evaluation, COVID risk index (CRI) of an individual. Figure 1 shows the risk curves of people with age for different comorbidities in a normalized scale of 100.

**Figure 1.**
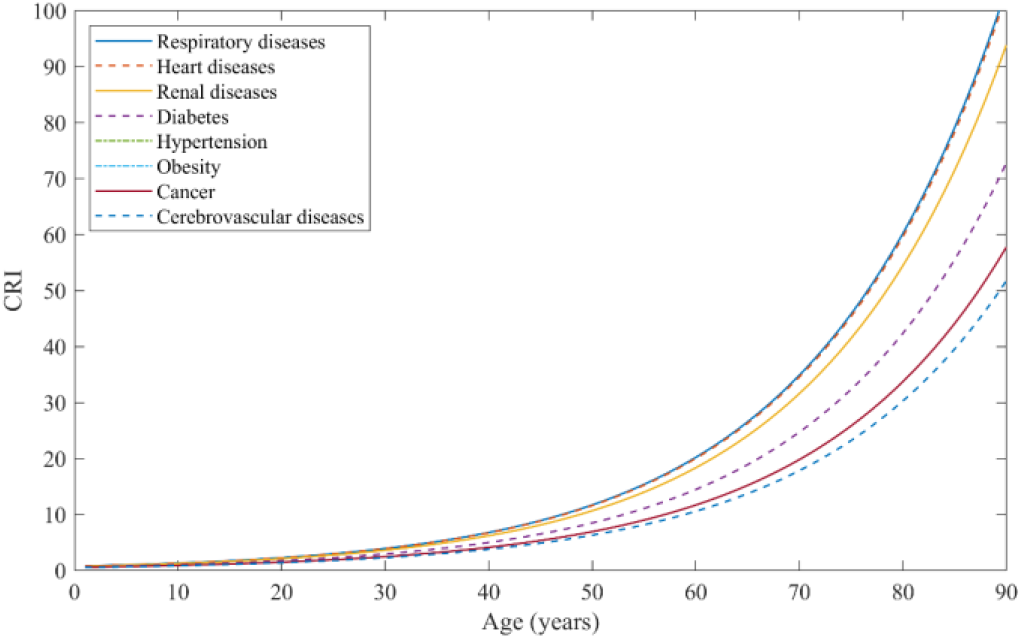
COVID risk index for men in relation with age and different comorbidities.

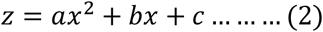

Where, coefficient values (with 95% confidence level) are, a = −3.40646579722815×10E-5 (−4.477e-05, − 2.336e-05), b = 0.00672965343817772 (0.00563, 0.007829), and c =0.635624565570039 (0.6123, 0.6589); with R-square value of 0.9994.

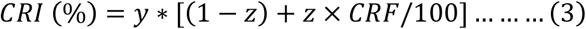

Risk is communicated as a five-point Likert scale to express very high, high, moderate, low and very low risk for an individual. Stratification of risk into five ranges is based on the distribution of the area under the risk curve (Figure 2). Where risk under the curve with maximum risk is distributed equally. Table 2 shows the range of CRI to represent a different level of risk.

**Table 2.** Qualitative representation of risk from CRI values.

| Qualitative risk level | CRI value range | Color code |
| --- | --- | --- |
| Very Low | $0 \leq \text{CRI} \leq 6$ | Dark Green |
| Low | $6 < \text{CRI} \leq 15$ | Light Green |
| Moderate | $15 < \text{CRI} \leq 28$ | Yellow |
| High | $28 < \text{CRI} \leq 48$ | Orange |
| Very High | $48 < \text{CRI} \leq 100$ | Red |

**Figure 2.**
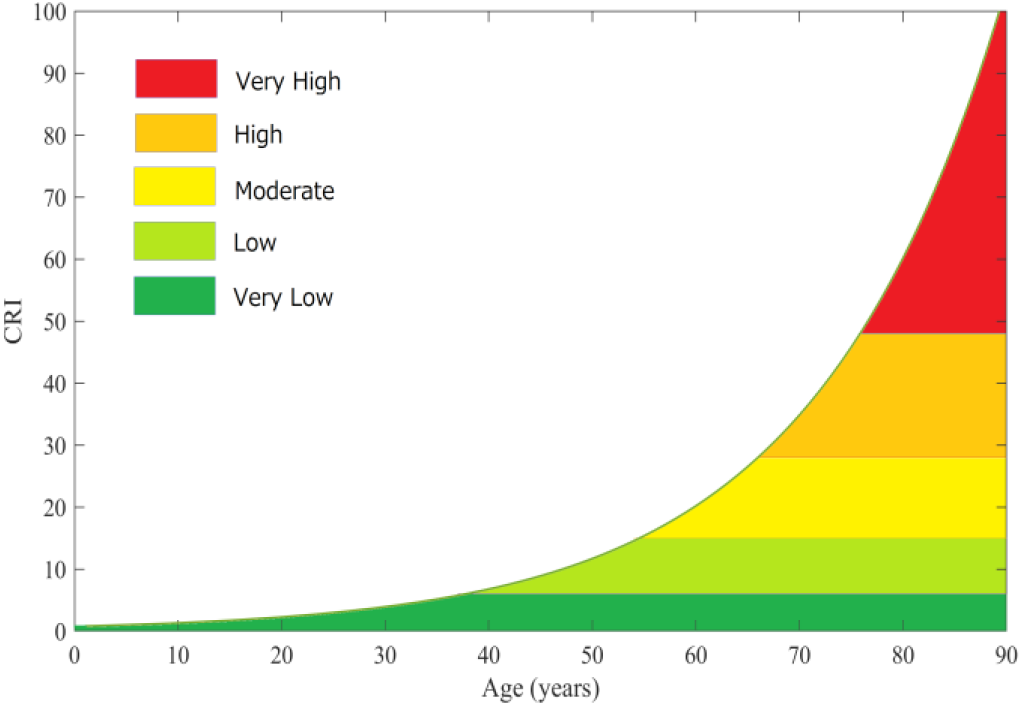
Risk level stratification considering equal risk area distribution over ranges.

#### Probability of COVID infection for individual

Probability of COVID-19 infection has been assessied using exposure and local risk. A detail explanation to calculate the exposure is discussed in the methodology. Probability of infection has been shown in a five risk-levels on a linear scale (0-20-40-60-80-100). Assessing the symptoms of the patients reported in recent studies[21]–[23], risk of having a positive case of COVID-19 in an individual is evaluated through their response to the questionnaire in a web app. Fever, cough and shortness of breath are major symptoms found in patients. Symptoms are only used to inform the respondent to contact to the nearest health care facilities depending on the level of their risks associated with symptoms.

### Regional risk assessment

Risk zones have been mapped through a multidisciplinary approach to evaluate the regional risk, which is applicable for any region across the world. Public health risk, considering the underlying health condition and health service facilities in respective of the total population of that area are mapped along with socioeconomic risk level associated with COVID-19 as shown in Figure 3. The overall scenario of COVID-19 risk and transmission risk throughout the country, considering the influential factors are presented for pre-pandemic scenario. Figure 4 shows the relative risk maps of Nepal for overall risk of COVID-19 and the transmission risk of COVID-19 on the pre-pandemic state.

**Figure 3.**
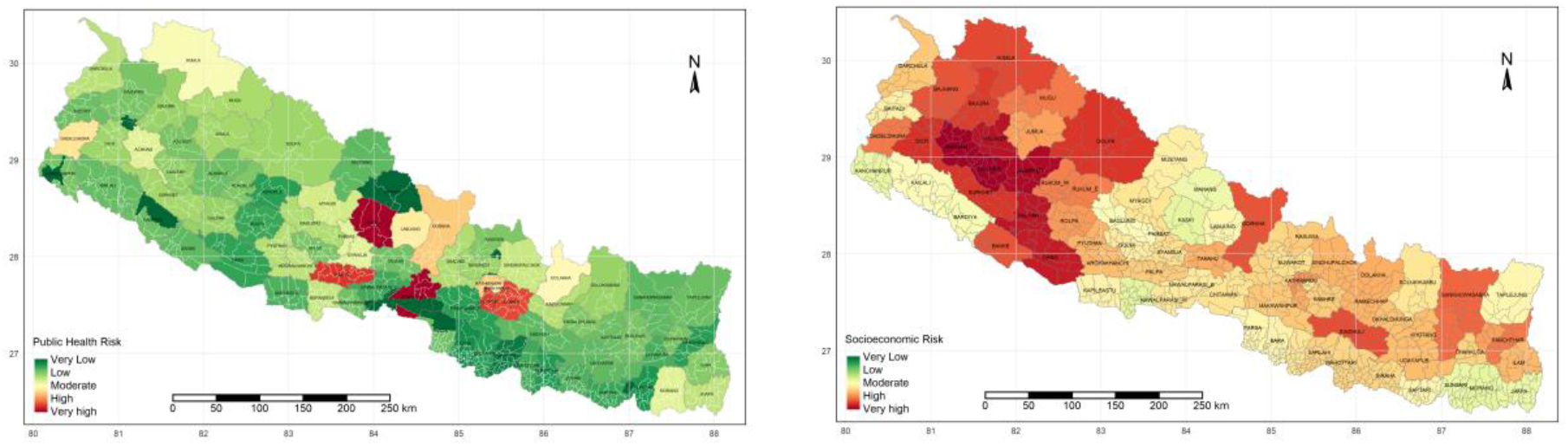
Public health risk (in left) and socioeconomic risk (in right) of Nepal for COVID-19 pandemic.

**Figure 4.**
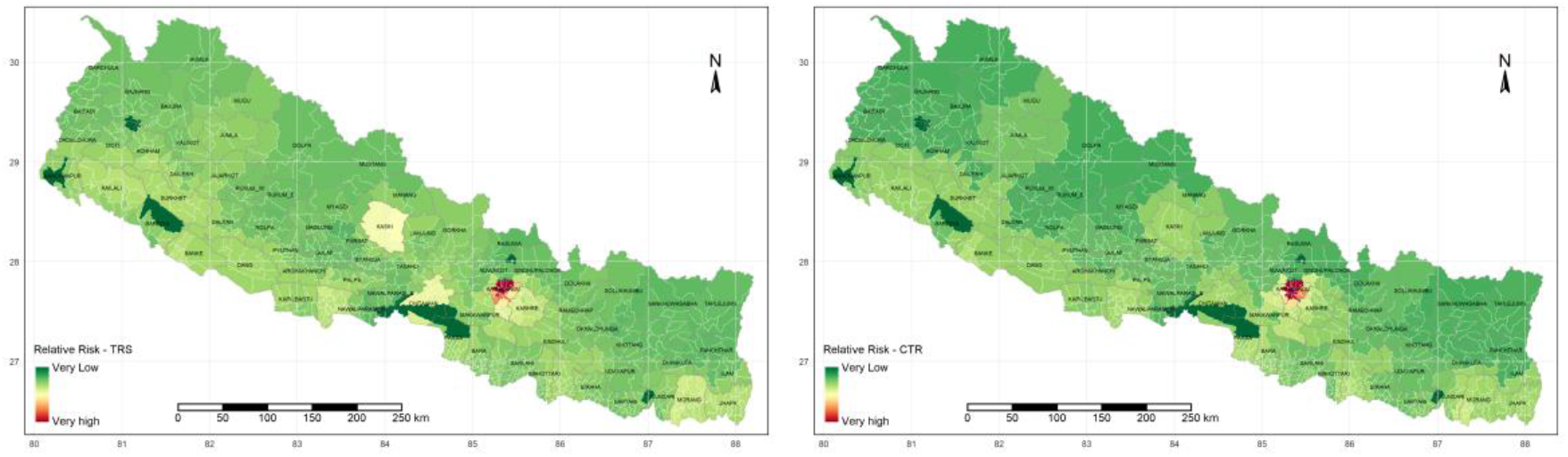
Relative risk maps of overall risk scenario, overall risk score of Nepal (Left) and COVID-19 transmission risk (CTR) (Right) prior to pandemic.

Kathmandu is in the high risk zone for both transmission and overall risk, however, most of the other areas with higher transmission risk are not associated with higher overall risk. Population exposure in Kathmandu is also highest in the country, hence the government should prioritize and localize the measures in the capital. CTR in southern boundaries are higher as it shares the open border with India, other areas with highly dense population, having more locations of exposure such as hotels, airports are in the upper side of the table.

Figure 5 shows the timeline of overall risk and CTR across the country on May 10^th^, 20^th^, 30^th^ and June 10^th^ respectively. This can be updated regularly, upon receiving the official data. It is presented in a continuous scale range from Very High to Very low-risk zones.

**Figure 5.**
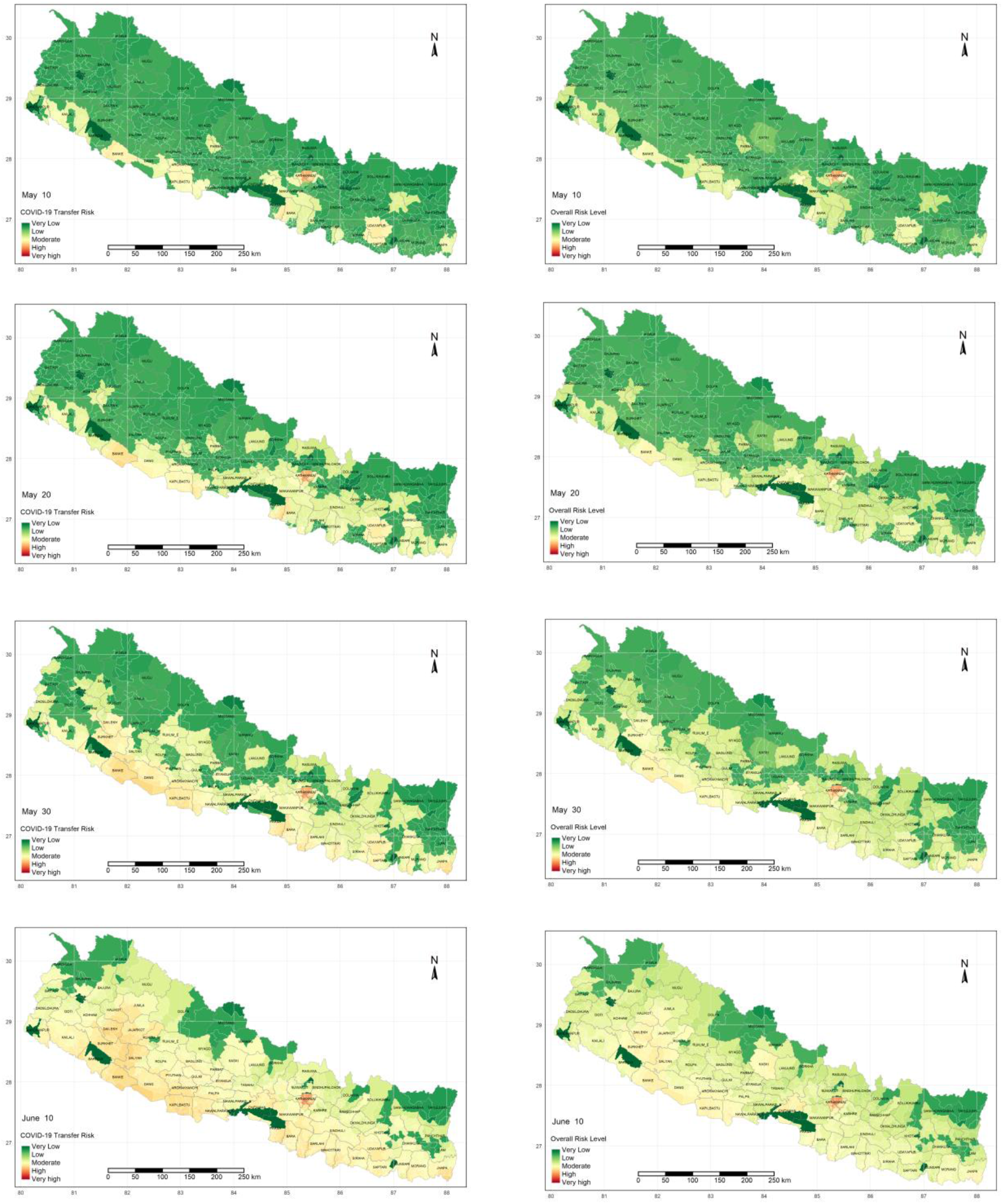
COVID 19 overall risk (in the left column) and CTR (right), updated after the positive cases confirmed May 10, 20 & 30 and June 10 from top to bottom.

Regional importance zonation is mainly based on food (rice and maize) production and supply chain across the region, Figure 6 shows the regional importance map of Nepal.

**Figure 6.**
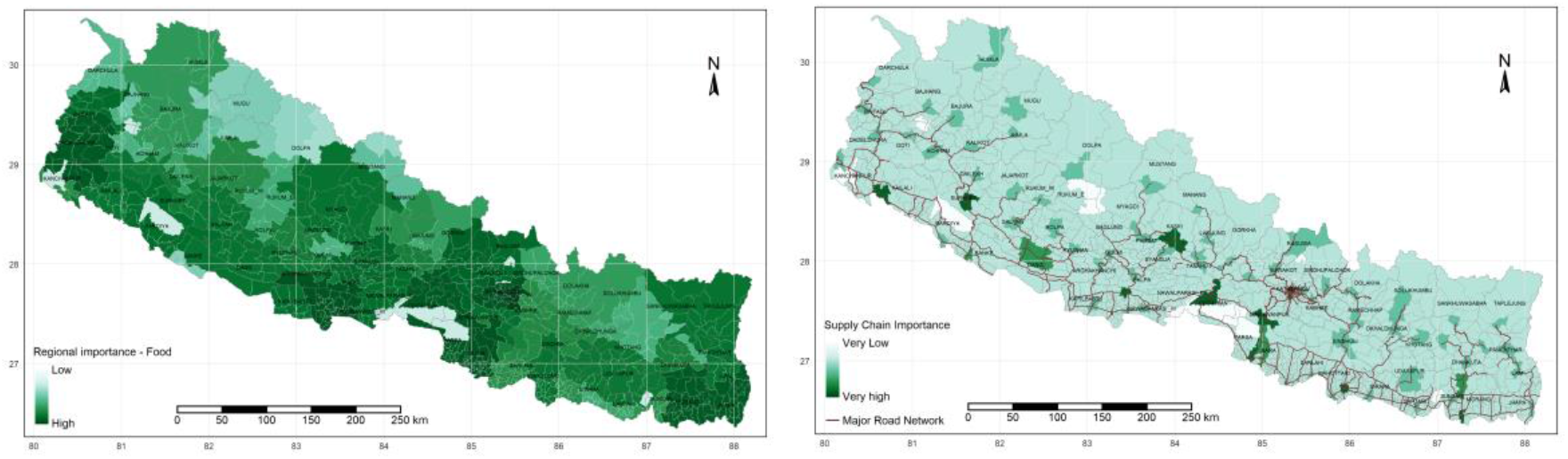
Regional importance food production (Left) & supply-chain importance (Right)

### COVIRA for risk assessment and communication

-COVIRA (www.covira.info) was launched by the Nepal Engineers Association on 21^st^ June 2020. Results are based on of data provided by the user questionnaire, designed for personal risk assessment. Address, age, existing health condition, any current symptoms, exposure to infected persons were collected. Data entered by an individual through multiple checkboxes are converted into scores which are then used to calculate risk using the equations above. Personal risk values solely depend on the data provided by the individual, but regional risk assessment, CTR will be the result of the risk in their locality. Result will be provided on the web as shown in Figure 7. Personalized messages are displayed in brief on information about risk and mitigation measures. Users can find the local risk in selected municipalities through the navigation on the page.

**Figure 7.**
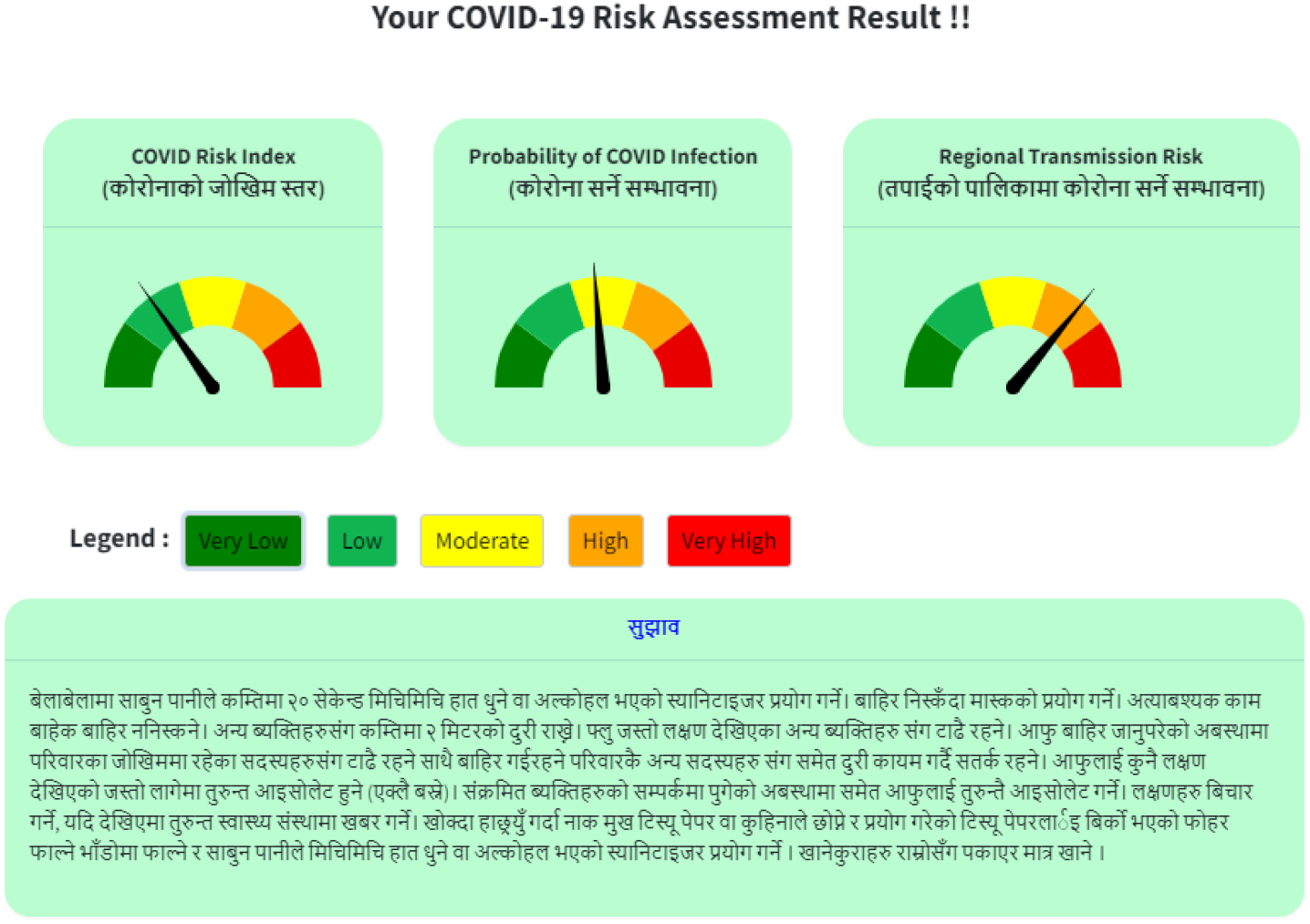
Result of COVIRA tool on-screen after personal risk assessment (in Nepali)

## Discussion

COVIRA framework would pave the way to move forward by minimizing the risk with no compromise in public health. Results from the model showed good resemblance of COVID-19 transmission risk when compared with no cases (right in Figure 4) and transmission risk on June 10^th^ with cases over the country (bottom left Figure 5). Hence, the cases are arising in the region where relative risks are higher and spreading over the lower-risk zones.

For regions with very low risk, where no active cases have been found, a vigilance placed at the community level can cautiously ease life from the nationwide lockdown. Regional importance and zoning of several factors to sustain the minimum level of public life for the future, such as food and other essentials production and supply, would guide the community to act responsibly. The use of this tool will also greatly assist in easing tight lockdowns in places where mass testing is not. A risk assessment using the tool will show those vulnerable in the region. It can also facilitate the entry of individuals into new zones after mobility is restored in countries, based on knowledge of population at risk.

The COVIRA currently developed for Nepal generates bothpersonal and zonal risk with a current scenario for adequate measurement and this tool can be scaled up worldwide. The two indicators for personal risk reflect the chance of transmission, infection, and recovery under the stipulated environmental and health condition. If the risk of transmission is high, one can limit one’s interactions with other people, which reduces the exposure. However, if one has likely to have a low life-threatening risk even if anyone gets infected with the COVID-19 they can continue their regular job with necessary precautions. This could help the people remain aware of based on their risk factors. The system generates the individual risk level along with public health advice to remain safe (see Figure 7). This way, a community level risk is also possible to get once a large number of people participate in this system.

Quantitative risk assessment is a challenging process, but a good method for communicating and making people aware of their vulnerability and risk due to COVID-19[26]. There is always a risk of entering the wrong information in a crowd-sourced data collection tool. However, if someone enters the wrong information and completes the assessment, it would only affect on their risk card since rRegional risk assessment relies on the depth of data available at community level.

The regional/zonal vulnerability risk is dynamic as the input variables are not static as these include, among others, the number of immigrants or COVID-19 positive cases. The COVIRA updates the vulnerability condition as the input parameters change. Figure 5, (a) represents the base vulnerability risk map, (pre-COVID-19), and Figure 5(d) is the latest vulnerability maps based on the updated COVID infected cases and number of immigrants quarantined from abroad. Specifically, a large number of people arrived from India after lockdown started and the majority of of newly identified COVID-19 infected people in this population. The base vulnerability map depicts that the urban area/region with international connections and regions bordering India have the highest base risk. And the total number of infected people has increased significantly in these high-risk regions in the base risk map.

A recent study in Nepal shows “three in every five employees lost their jobs due to the COVID-19”, and the resulting shardship could be even worst next year if total lockdown is continued[27]. Therefore, the personal risk and zonal risk map can help some people to continue their daily activities, based on its priority and risks. On March 24, when the Nepal government imposed the nationwide lockdown, there was only one active COVID-19 patient and no casualty. Instead of taking a local approach, the government imposed a nationwide absolute lockdown as its blanket approach, perhaps alocal approach of containing the virus could have implemented for a better balance between health and socio-economic consequences.

The scenario could change rapidly based on the infected condition, local activity, and the immigration condition, data availability on time is always an issue. The reference data are taken from the high-income countries (Italy, Spain, Germany, USA, UK) and China which are different from LMICs countries like Nepal. However, the varying data sets of those countries well represent the risk scenario all over the world.

## Methodology

Risk assessment is principally based on the available data, risk assessment in pandemics mainly rely on daily updates on infected cases and recoveries. Main three sources of data are used; 1) recent updates about COVID-19 (to find the relationships between potentially influential factors and to update the risk map); 2) demography of health condition and facilities, socioeconomic and other static data; and 3) questionnaire response in the app of an individual to assess the personal risk.

### Data sources

Published data related to COVID-19 cases from China[28] (n=1023), Italy[28] (n=1624), Spain[29] (n=16680), Germany[30] (n=6512), the USA[31] (n=12998) and the UK[25] (n=20483) are used to establish the relationships of different factors associated with the virus spreading and loss of life. The individual risk depends upon age and underlying health conditions, however, other contextual demographic classifications, as like ethnicity may further extend in different countries. Regional risk assessment for most of the pandemic depends on population with an underlying health conditions, health infrastructure facilities and services, water sanitation and hygiene status, poverty, literacy, population density etc[32]. This study provides an example of regional risk assessment for Nepal, using the data published in national reports[33]–[35]. Daily updates on COVID cases and quarantines are linked with the Ministry of Health and Population web[12].

#### Personal risk assessment

Personal risk assessment consists of major three parts: 1) individual vulnerability (danger to life when infected with COVID-19); 2) risk of infection if symptoms exist and 3) risk of transmission to individuals are being evaluated from their questionnaire responses.

Age and existing health conditions are two key factors for individual vulnerability. Distribution of death of different age groups is further converted in relative scale from different countries. Death toll distribution over the age group from the total death in the country and corresponding positive COVID cases of the age group are analyzed. Figure 8 shows the normalized fatality rates for different age groups from two perspectives; first column (a, b and c) shows the fatality rates in reported positive cases for each age group where the second (d, e and f) represent relative death percentage of each age group in totality. Interestingly, relative death percentages of age group to the total death and relative death percentage to the confirmed cases of the corresponding age group show a very good correlation irrespective of sample size and country except for China. Data from China show a lower percentage of death to total death in the highest age group, however, the ratio of death to a positive case is highest in that age group. Equation 1 has been derived from death to positive cases percentage in age groups to represent the COVID-19 risk factor.

**Figure 8.**
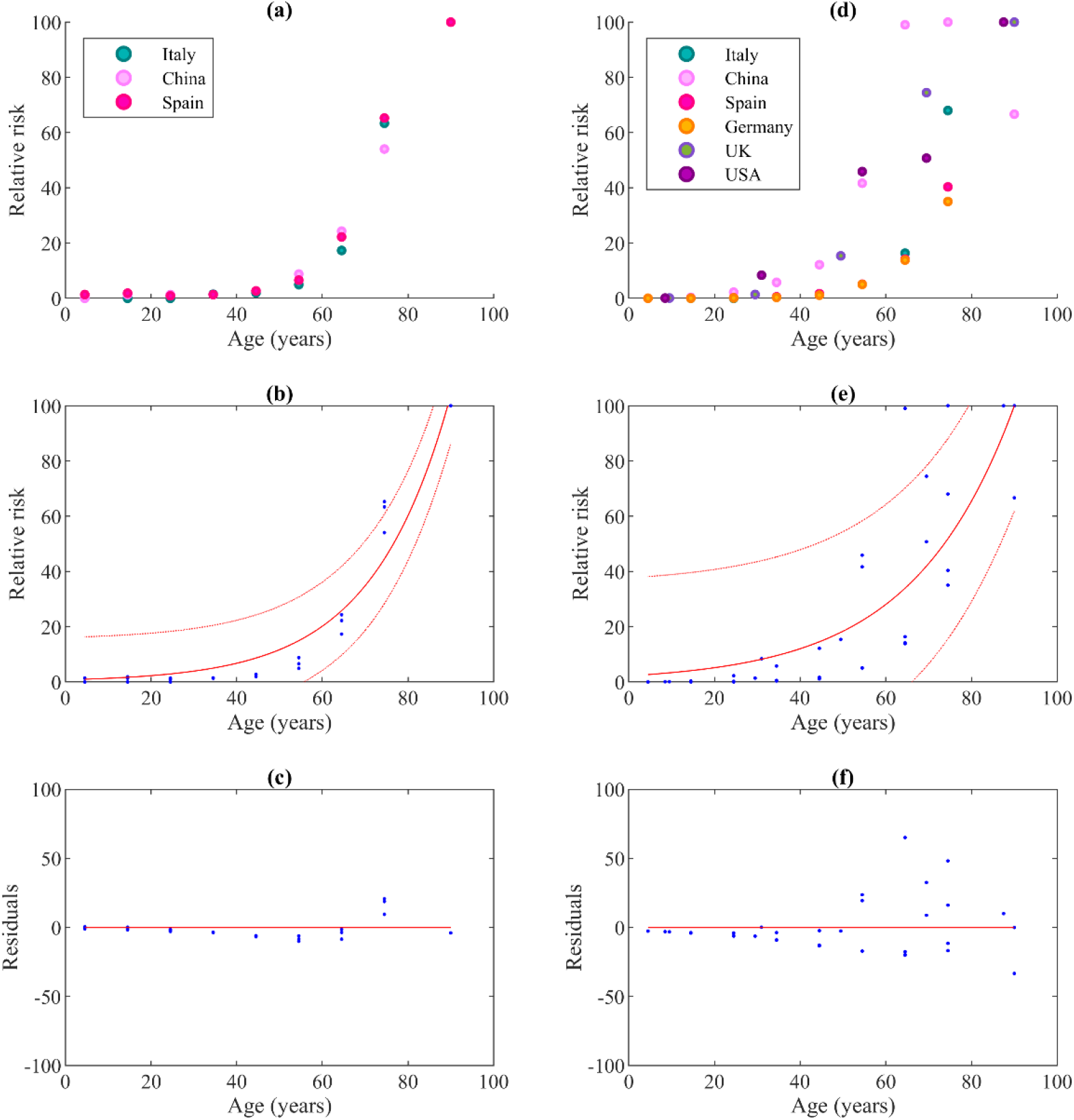
Correlation of recorded deaths and age; a) normalized fatality percentages of cases of respective age group; b) exponential curve fit to data of ‘a’ with 95% confidence boundaries and c) corresponding residuals, d) normalized fatality percentage of total fatality reported, e) exponential curve fit to data of ‘d’ with 95% confidence boundaries and f) corresponding residuals.

Underlying health condition of the reported cases is also analyzed using the data those reported from China[21], [22], the USA[23], and the UK[24]. Four sets of data are used to find the relative risk of different existing health conditions, even though all data sets are not in a similar format, they support each other in away. The major risk factor was established from the percentage of death and or severe condition of patients with different underlying health conditions. Obesity has not been listed in data from China but the USA and UK both have large numbers of patients even though the death percentage is not clearly mentioned but is in nearly the same range of hypertension. Hence, the obesity risk factor is assigned similar to hypertension. The chronology of data tables and calculations are provided in the annex. The average of relative risk factors of different existing health conditions are provided in Table 1. A recent study on gender effect on mortality rates due to COVID-19 suggested the higher mortality rate for men (70.3% vs 29.7%)[36]. Hence, having the risk factor for male is 100, maximum risk factor for female will be 42.24. Total risk evaluated will be multiplied by 0.4224 for female.

Individual vulnerability is now calculated through the weightage of age, gender and existing health conditions. Recent data from the USA[23] and the UK[25](as of April 31) show the death percentages of COVID cases with and without underlying health conditions are 89.3% (n=1,482) and 94.8% (n=19,740). Following the trend of the percentage of deaths with an existing health condition in different age groups, another function for the coefficient of comorbidity has been derived from UK data as previously shown in equation 2.

#### Probability of COVID-19 infection for individual

Table 3 provides the relative percentage of patients having different symptoms from the data provided in referred studies. Fever is the most common symptom where cough is almost found in a similar number of patients, shortness of breath, fatigue, and sputum production are other major symptoms. Considering the asymptomatic cases of COVID-19, providing the probability of having COVID-19 result is challenging. Hence, if someone report any one symptom or combination of different sysmptoms, recommendations will be displayed to contact to the nearest health care facilities.

**Table 3.** Symptoms and corresponding relative pecentages.

| Symptoms | Relative percentage |
| --- | --- |
| Fever | 100 |
| cough | 87 |
| Shortness of breathe | 57 |
| Fatigue | 34 |
| Sputum production | 31 |
| Myalgia | 24 |
| Sore throat | 18 |
| Headache | 17 |
| Chest pain | 17 |
| Diarrhea | 14 |
| Nausea or vomiting | 13 |
| Chills | 13 |
| Nasal congestion | 12 |

In addition, exposure of any individual can be calculated through their profession, daily activities, travel, and meeting with (a-symptomatic) COVID-19 infected people. Exposure and corresponding values are shown in table in appendix. Now, considering the exposure and regional risk, the total probability of COVID-19 infection risk will be calculated. Initial phase of Nepal cases as reported in media briefings by Ministry of Health, very few of the people having positive case have symptoms. Hence, we proposed the risk of COVID-19 infection as a product of local risk and exposure. Equation 4 provides the probability of infection, where the upper bound value will be limited to 100, even if it exceeds.

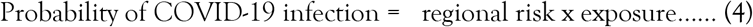

Exposure can be evaluated through the occupation, proximity to public and other persons and their day to day safety measures. Studies from Italy[37], and six Asian countries[38] suggested the risk levels for different work groups, where drivers and health workers have an higher exposure. Exposure level in scale of 0 – 100 are provided in Table 4 and Table 5 on the basis of these recent studies.

**Table 4.**
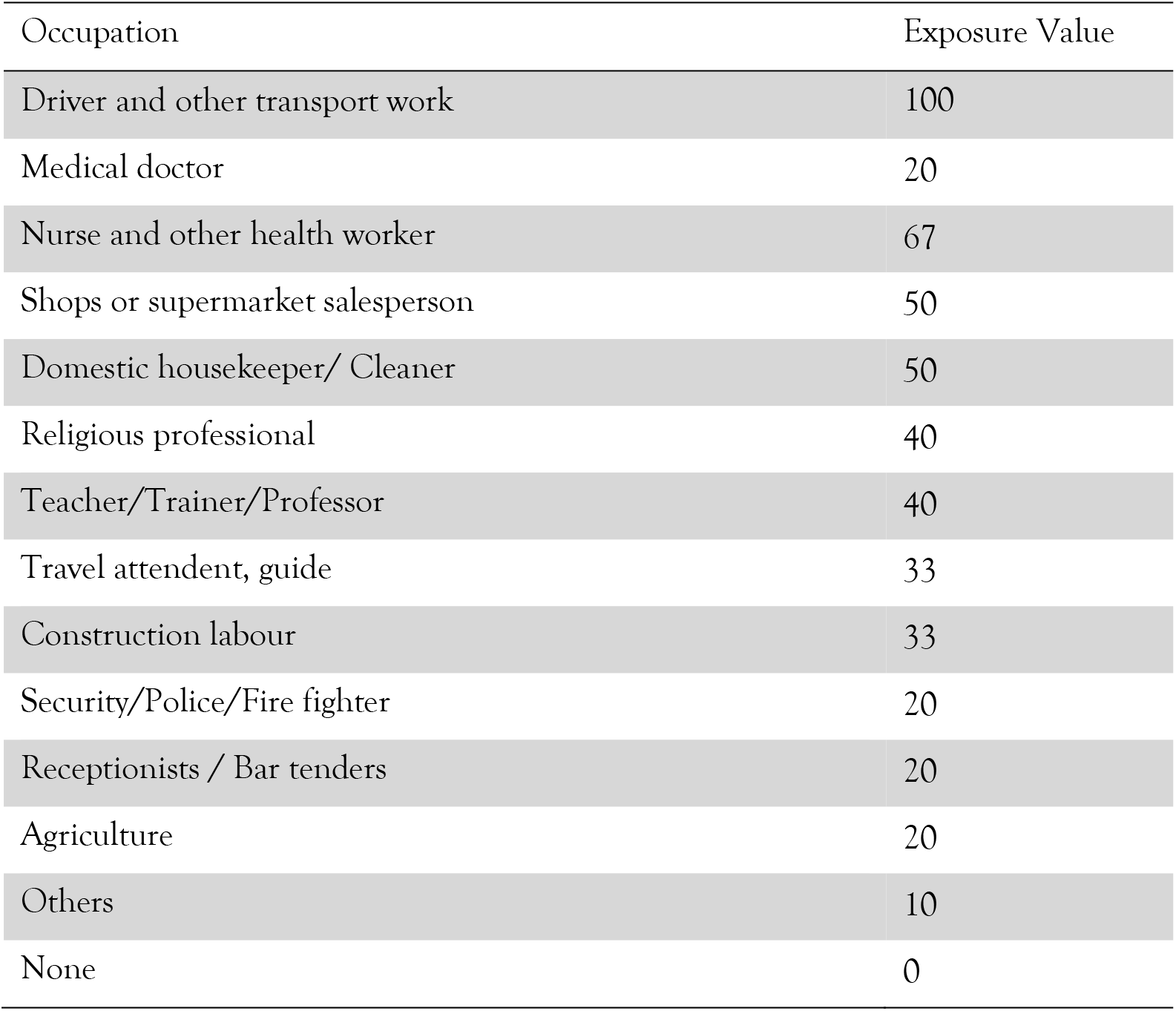
Occupation and exposure value.

**Table 5.** Proximity and exposure value.

| Activities | Exposure Value |
| --- | --- |
| Travelled to COVID-19 infected area recently | 100 |
| Met with known COVID19 infected person | 100 |
| Have to be in proximity of less than 2 meters in workplace (or in field, as part of job) with public | 90 |
| Need to travel/walk in public areas where maintaining the distance of more than 2 meters is impossible. | 80 |
| Need to attend meetings in person occasionally | 70 |
| Have to be in proximity of less than 2 meters in workplace with other colleagues | 60 |
| Have more than one family members, who need to go out and meet other people | 50 |
| Have family member, who need to go out and meet other people | 25 |
| None | 0 |

In addition, preventive measures including using of face masks, keeping physical distances, wearing sun glasses in daily life reduce exposure and hence risk[39]. Hence, contribution in reduction of exposure has been introduced as shown in Table 6 those on the basis of results presented by Chu et al.[39]. Differences in risk presented due to each preventive measures are directly used to reduce the exposure level. Regular sanitization has also reduce the exposure, hence considering the minimum exposure level (50); with all preventive measures, net exposure is nullified.

**Table 6.** Preventive measures and deduction in exposure value.

| Preventive measures | Exposure Value |
| --- | --- |
| Wear mask when going to crowded areas | -14 |
| Sanitize hands using hand washing or sanitizer | -15 |
| maintain physical distance when going outside | -10 |
| use sunglasses/face shields | -11 |

#### Regional risk assessment

A multidisciplinary framework to evaluate the regional risk has been proposed to provide support on risk zoning. COVID-19 risk can be stratified into different levels based on COVID-19 transmission risk, public health status and facilities and socioeconomic condition. Population with an underlying health condition and older people are more vulnerable[40], hence those regions with different existing health condition population are assigned score in a scale of 100. Available health facilities like hospitals, primary health care centre, health posts and female community health volunteers are also relatively scored in a scale of 100, throughout the country, in a proportion of the population.

Population with higher poverty levels are more vulnerable during the pandemic, loss of a job, shortage of food could push them further down in poverty, likely to have poor nutrition, and loosening the immunity power. Hence, the socio-economic index of the region has also contributed to the vulnerability of the region, where literacy rates and water, sanitation and hygiene indexes are also used to further clarify the risk scenario.

Regional food production capacity and type, major supply chain hubs and network are further categorized with relative indices. Government guidelines and policies along the pandemic period may vary additional parameters to consider in importance zoning. General components of the framework are shown in Figure 9. There is a lack of proper statistical data to calculate the factors for different components in the overall risk calculation related to this pandemic, hence, a Delphi method has been used to propose the equations [41], [42].

**Figure 9.**
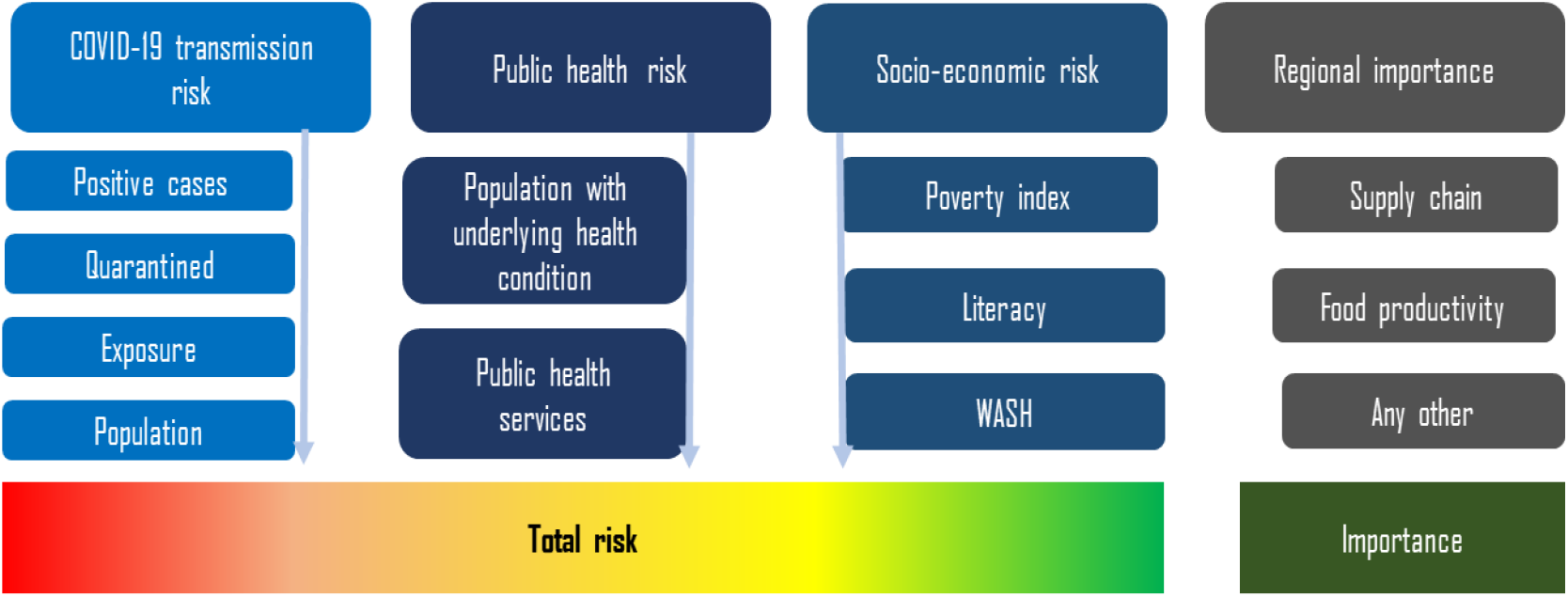
Framework and components for total COVID-19 risk evaluation.

##### COVID-19 transmission risk

Human to human transmission of the virus poses higher risk where more positive cases are found, other factors contributing to regional risk are, quarantined people in the region, who have visited or travelled from high-risk zones; exposure of the region to other regions of high risk and the population density. Equation 5 has been proposed for COVID-19 transmission risk.

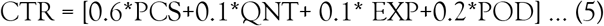

where, PCS = Positive case score

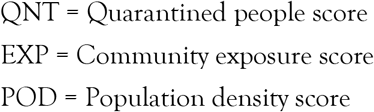

PCS is calculated in a relative scale of 0 to 100, considering the total active cases in and nearby region. Having zero cases, there will be no score of PCS but when the region witnessed any case then the risk score starts from 50. The reason behind the benchmark of ‘50’ is that, any active case may infect further cases in that community, hence assigned 50 for the first case, then risk will be increased in logarithmic scale to be 100 when cases will match with total household in the community. The score matrix of PCS for the region is calculated as shown in Equation (6). Figure 10 shows the scenario of PCS for number cases for different administrative regions [Kathmandu (985,453), Pokhara (426,759), Lalitpur (284,922), Machhapuchhre (21,868), Chame (1,129) and Narpa Bhumi (538)] with a maximum and minimum population in the country. The risk transmission possibility of the active cases from an administrative region to the nearby region is also considered. Most people from the boundary region could travel to the neighouring region for shopping, work or tradeas there are no restriction on movement. All adjacent regions of the index region having cases is considered the same active case to compute the PCS within the 10km buffer. It is considered half in the administrative regions within another 10km from the main region having active cases.

**Figure 10.**
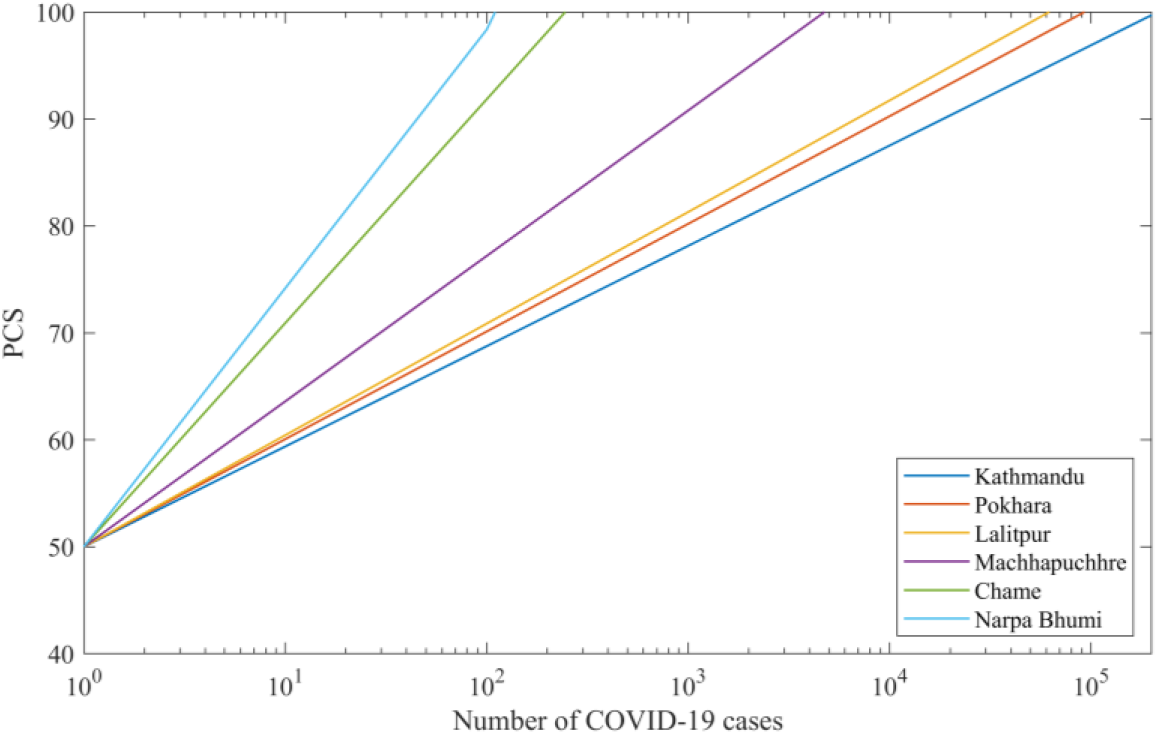
PCS for different urban/ rural administrative regions on number of cases.

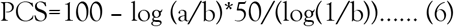

Where ‘a’ is the number of active cases in the region ‘b’ is a total number of household. Average household size of Nepal is 4.6[43], which is used to calculate the household numbers in each administrative unit. The risk will be minimized over time if no cases are found in the considered administrative zone. Considering the characteristics of COVID-19 as reported by Lauer etal., time of probable exposure to the detection of the positive case is up to 60 days for most of the cases, however very few are longer but less than 90 days[44]. Hence, Any administrative unit, considered in this study, having null positive cases for 60 consecutive days, PCS will be reduced to 50% of its value and after another consecutive 30 days (total 90 days) of no positive cases reports, PCS will be nullified.

The score for the quarantined people in the region follows the similar approach of the PCS. Exposure of the region is scored based on facilities those use by the people out of the region, like hotels, airports are considered. International arrivals include international airport and the other land and port immigration points. Exposure score is tabulated in Table 7 for different facilities in the region.

**Table 7.** Exposure values for the combination of different facilities.

| International arrivals | Domestic airport | 3+ star Hotels | Other hotels | EXP value |
| --- | --- | --- | --- | --- |
| √ | √ | √ | √ | 100 |
| √ | X | X | √ | 90 |
| X | √ | √ | √ | 80 |
| X | √ | X | √ | 70 |
| X | X | √ | √ | 60 |
| X | X | X | √ | 50 |

Population density of the region linearly correlates with the prevalence rate of infectious diseases[45]. Hence, the score for the population density of the region is relatively linearly distributed throughout the country with the minimum assigned score of 20.

#### Public health risk

People with different underlying health condition (UHC) and the low availability of health facilities (HF) in the region are associated to the public health risk (PHR) of that region. People with different UHC in the region are scored in a single value with the weightage factors as of Table 1. Hospitals, primary health care centers, and health posts are assigned 100,10, and 1 respectively for each unit. Summation of unit values in the region will be divided by the population in the region. Relative score on scale of 100 will be calculated for each region. Both UHC and PHF are in relative scale of 100. Hence, considering only 20% would potentially get service to recover from critical stage, PHR is given by the equation 7.

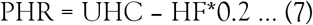

Further assessment of public health in community can be extended with different pandemic and post-disaster scenario. Natural hazard and structural vulnerability, production and supply of medical facilities and medicines along with managerial and social engagement can also be considered in the public health vulnerability. For the scope of COVID-19, this study confined within the smaller boundary.

#### Socioeconomic risk

Socioeconomic risk (SER) for the pandemic is also considered in the integrated risk analysis. Poverty not increases to the risk of hunger, but poor people are often not able to afford the required precautionary measures. Lower literacy rate indicates the lower level of awareness and similarly lower index of water sanitation and hygiene indicates the higher chances of spreading of the virus. Poverty index (PVI), literacy rate (LTR) and water, sanitation and hygiene index (WASH) of the region are relatively scored in scale of 100. Awareness campaigns through media and community can increase the public awareness for even simply literate people so it will be given lower factor, comparison to poverty and WASH which would not change within a short time frame. Combining the factors, SER can be represented by the equation 8.

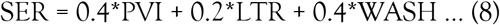

#### Regional importance

Food production and supply is the most important maintain general health and well-being in the community, which ultimately support to fight the pandemic. Hence, major hubs of supply chain network (SCN) are mapped for their importance in scale of 100 as shown in Table 8. Similarly, food productivity (FOOD) of the region are also relatively scored. Rice, wheat and maize production are taken into account with their planting and harvesting time which will be more crucial for that region. Additionally, supply of fertilizers, seeds and other supporting materials should be available within the very short time frame. We cannot postpone the farming of such crops by months or even weeks, it depends on the climate and weather. Food production and supply chain, both are equally important, hence, regional importance (REG) can be represented in separate maps.

**Table 8.** The score of major hub for supply chain network.

| Supply chain hubs | Score |
| --- | --- |
| Capital city | 100 |
| Provincial headquarters | 90 |
| Major cities/hubs other than above | 70 |
| District headquarters | 50 |
| Others | 30 |

Overall risk will be calculated by combining CTR, PHR and SER, on the other hand regional importance will be mapped. CTR is the major component in total risk score (TRS), where PHR and SER will further escalate the risk, as proposed in equation 9.

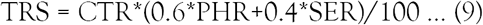

## Conclusion

COVIRA, a risk assessment and communication tool has been designed to provide an overall risk assessment framework for COVID-19. The tool has been tested for Nepal but can be easily adapted across the globe, since risk factors are similar worldwide. The empirical models fitted through the the available data sets of COVID-19 are used for evaluation of individual risk, considering major risk factors identified into the historical dataset. Personal risk assessment through questionnaire provides the individual risk and communicate the rules and guidelines for awareness. Regional risk assessment would support government and community to act locally, which will give a clear way out to manage the risk wisely, refrained from any compromise on public health. Specifically, the regional risk and transmission map could help support essential supply chains at the time of absolute lockdown across the country. This multidisciplinary, holistic approach of risk assessment and communication can be extended to any other future pandemic and natural disasters.

## Data Availability

All data available within the paper

## Data Availability Statement

All data and models generated or used during the study appear in the submitted article including supplementary materials.

## Acknowledgement

First author would like to thank Prof. Anastasios Sextos and SAFER project team members in University of Bristol for their comments and suggestions. Authors are grateful to Nepal Engineers Association and Chairman Prof. Triratna Bajracharya, for endorsing the tool in Nepal after brief review. Authors would like to thank Mr Anil Pokhrel, Chief Executive at National Disaster Risk Reduction and Management Authority (NDRRMA), Nepal for his comments and suggestions. Authors would extend their thank Bipin Khatiwada and Sujan Parajuli for their support on web development and data update.

## Contributions

R.R.P. conceptualized, designed and drafted the paper; B.M. analyzed spatial data and produced most of the figures; R.R.P analyzed the statistical data, B.M., A.B, B.R.G, S.P., K.S., S.M.D., S.K.S. involved in drafting significant proportion of the paper or figures; P.S. and E.V.T. critically revised the paper.

## Appendix-A

### Data for proportion of death under each age group in total death

**Table A-1.**
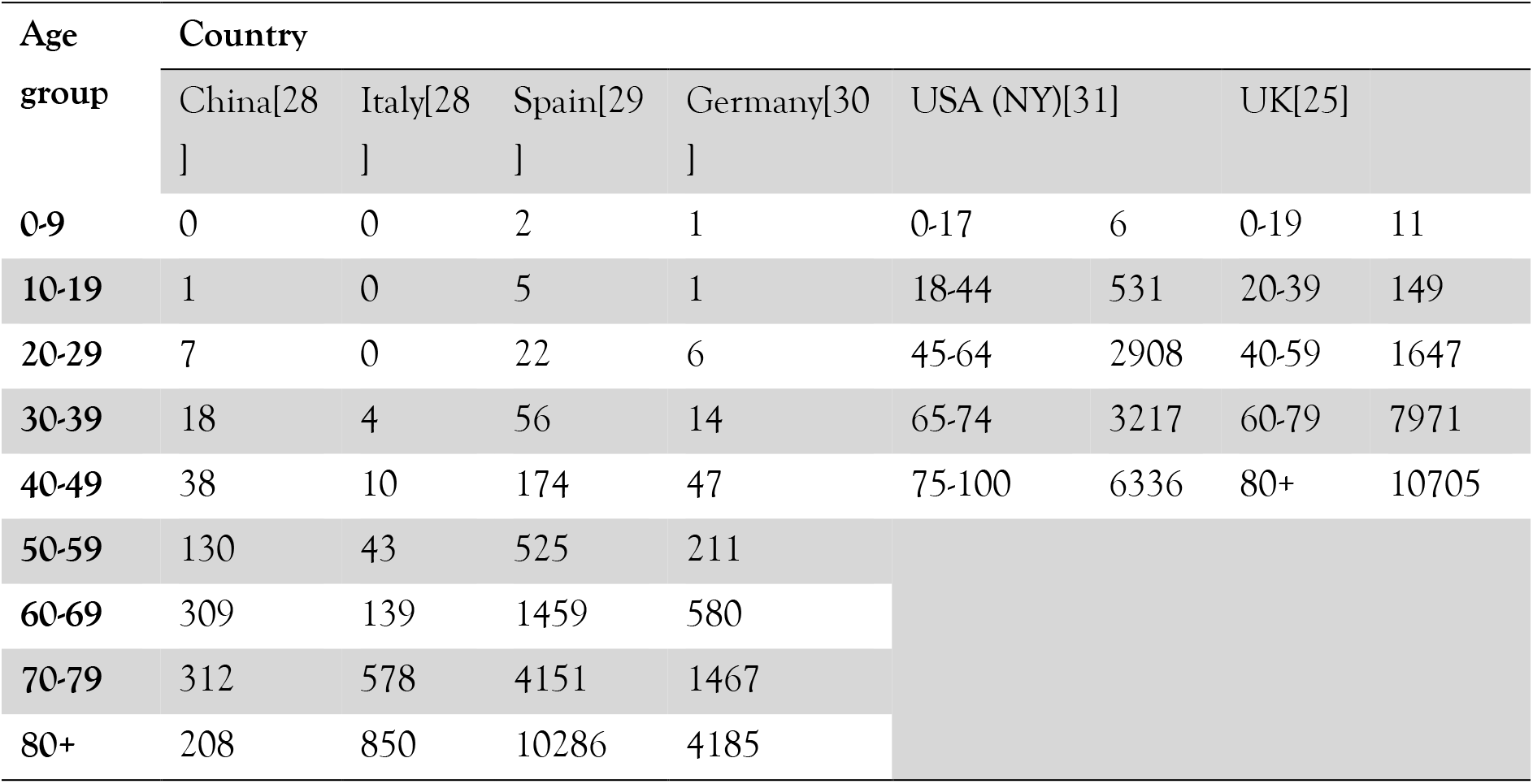
Death cases reported under each age group (Numbers)

**Table A-2.**
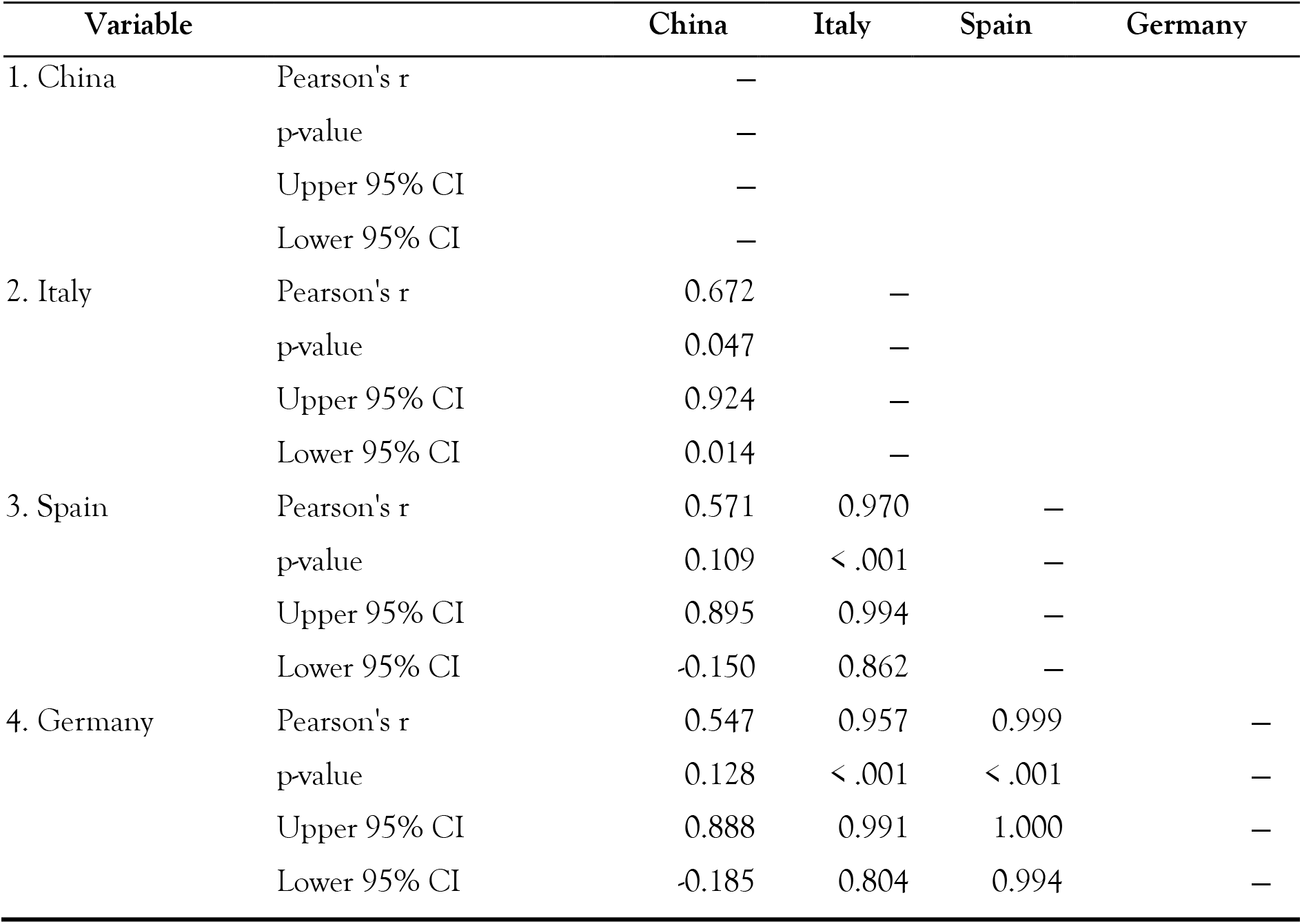
Pearson’s Correlations.

| Variable |  | China | Italy | Spain | Germany |
| --- | --- | --- | --- | --- | --- |
| 1. China | Pearson's r | — |  |  |  |
|  | p-value | — |  |  |  |
|  | Upper 95% CI | — |  |  |  |
|  | Lower 95% CI | — |  |  |  |
| 2. Italy | Pearson's r | 0.672 | — |  |  |
|  | p-value | 0.047 | — |  |  |
|  | Upper 95% CI | 0.924 | — |  |  |
|  | Lower 95% CI | 0.014 | — |  |  |
| 3. Spain | Pearson's r | 0.571 | 0.970 | — |  |
|  | p-value | 0.109 | < .001 | — |  |
|  | Upper 95% CI | 0.895 | 0.994 | — |  |
|  | Lower 95% CI | -0.150 | 0.862 | — |  |
| 4. Germany | Pearson's r | 0.547 | 0.957 | 0.999 | — |
|  | p-value | 0.128 | < .001 | < .001 | — |
|  | Upper 95% CI | 0.888 | 0.991 | 1.000 | — |
|  | Lower 95% CI | -0.185 | 0.804 | 0.994 | — |

**Table A-3.**
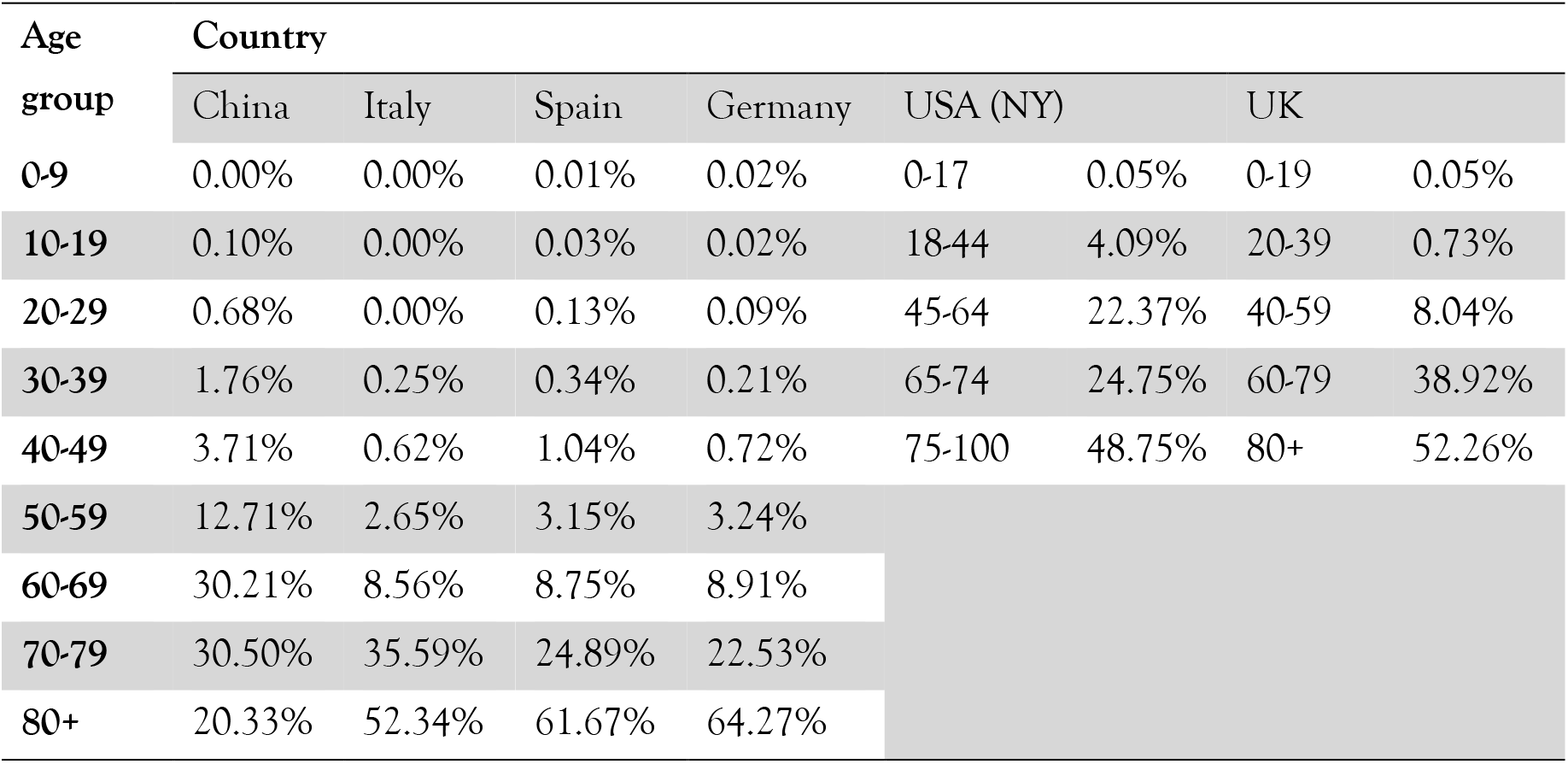
Death cases reported under each age group (percentage)

### Data for proportion of death under each age group in total cases of that age group

**Table A-4.**
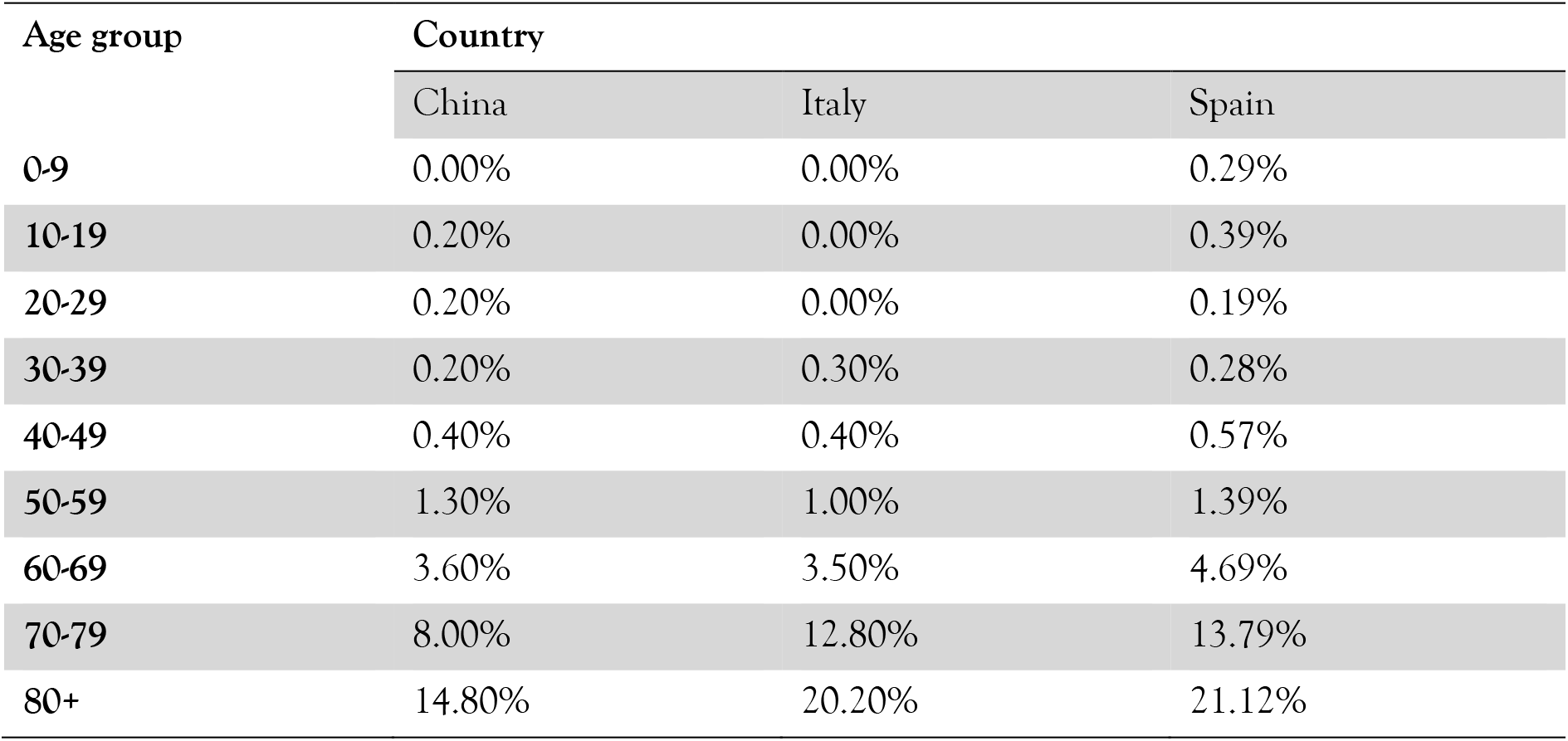
Fatality percentage of confirmed cases under each age group.

**Table A-5.** Pearson’s Correlations.

| Variable |  | China | Italy | Spain |
| --- | --- | --- | --- | --- |
| 1. China | Pearson's r | — |  |  |
|  | p-value | — |  |  |
|  | Upper 95% CI | — |  |  |
|  | Lower 95% CI | — |  |  |
| 2. Italy | Pearson's r | 0.993 | — |  |
|  | p-value | < .001 | — |  |
|  | Upper 95% CI | 0.999 | — |  |
|  | Lower 95% CI | 0.968 | — |  |
| 3. Spain | Pearson's r | 0.994 | 0.999 | — |
|  | p-value | < .001 | < .001 | — |
|  | Upper 95% CI | 0.999 | 1.000 | — |
|  | Lower 95% CI | 0.972 | 0.996 | — |

### Calculation of health condition risk factor

**Table A-6.** Percentage of patients with different underlying health conditions.

| Source | Guan et al<sup>6</sup> |  | Zhou et al<sup>7</sup> |  | Average of both data (in %) | Relative risk factor |
| --- | --- | --- | --- | --- | --- | --- |
| Comorbidities | Total cases | Severe cases | Total cases | Death cases |  |  |
| Chronic obstructive pulmonary disease | 12 | 6 | 6 | 4 | 56% | 100 |
| Diabetes | 81 | 28 | 36 | 17 | 38% | 69 |
| Hypertension | 165 | 41 | 58 | 26 | 30% | 54 |
| Cardiovascular/Coronary heart | 27 | 10 | 15 | 13 | 55% | 99 |
| Cerebrovascular | 15 | 4 |  |  | 27% | 48 |
| Cancer | 10 | 3 |  |  | 30% | 54 |
| Renal | 8 | 3 | 2 | 2 | 50% | 90 |

**Table A-7.** Death cases of UK, with and without existing health conditions (data of April 31)

| Age | with | without | Total | % with health condition |
| --- | --- | --- | --- | --- |
| 0-19 | 7 | 3 | 10 | 70% |
| 20-39 | 117 | 28 | 145 | 81% |
| 40-59 | 1401 | 184 | 1585 | 88% |
| 60-79 | 7270 | 433 | 7703 | 94% |
| 80+ | 9925 | 372 | 10297 | 96% |

**Table A-8.** Percentage of patients with different symptoms.

| Source | Guan et al[21] | Zhou et al[22] | Garg et al[23] |
| --- | --- | --- | --- |
| Country | China | China (Wuhan) | USA |
| Sample | 1099 | 191 | 180(Max) |
| Fever | 88.7 | 94 | 85 |
| Nasal congestion | 4.8 |  | 16.1 |
| Headache | 13.6 |  | 16.1 |
| cough | 67.8 | 79 | 86.1 |
| Sore throat | 13.9 |  | 17.8 |
| Sputum production | 33.7 | 23 |  |
| Fatigue | 38.1 | 23 |  |
| Shortness of breathe | 18.7 |  | 80 |
| Nausea or vomiting | 5 | 4 | 24.4 |
| Diarrhea | 3.8 | 5 | 26.7 |
| Myalgia | 14.9 | 15 | 34.4 |
| chills | 11.5 |  |  |
| Chest pain |  |  | 15 |

**Table A-9.** Relative percentage of patients with different symptoms.

| Source | Guan et al<sup>6</sup> | Zhou et al<sup>7</sup> | Garg et al<sup>8</sup> | Average |
| --- | --- | --- | --- | --- |
| Country | China | China (Wuhan) | USA |  |
| Sample | 1099 | 191 | 180(Max) |  |
| Fever | 100% | 100% | 99% | 100% |
| Nasal congestion | 5% |  | 19% | 12% |
| Headache | 15% |  | 19% | 17% |
| cough | 76% | 84% | 100% | 87% |
| Sore throat | 16% |  | 21% | 18% |
| Sputum production | 38% | 24% |  | 31% |
| Fatigue | 43% | 24% |  | 34% |
| Shortness of breathe | 21% |  | 93% | 57% |
| Nausea or vomiting | 6% | 4% | 28% | 13% |
| Diarrhea | 4% | 5% | 31% | 14% |
| Myalgia | 17% | 16% | 40% | 24% |
| chills | 13% |  |  | 13% |
| Chest pain |  |  | 17% | 17% |

## References

[1] World Health Organization, “Situation Report,” 2020.

[2] F. Wu et al., “A new coronavirus associated with human respiratory disease in China,” Nature, vol. 579, no. 7798, pp. 265–269, 2020.

[3] M. A. Shereen, S. Khan, A. Kazmi, N. Bashir, and R. Siddique, “COVID-19 infection: Origin, transmission, and characteristics of human coronaviruses,” J. Adv. Res., vol. 24, pp. 91–98, 2020.

[4] C. Sohrabi et al., “World Health Organization declares global emergency: A review of the 2019 novel coronavirus (COVID-19),” Int. J. Surg., vol. 76, no. February, pp. 71–76, 2020.

[5] R. M. Piryani, S. Piryani, and J. N. Shah, “Nepal’s Response to Contain COVID-19 Infection.,” J. Nepal Health Res. Counc., vol. 18, no. 1, pp. 128–134, Apr. 2020.

[6] WHO, “Responding to community spread of COVID-19,” 2020.

[7] J. Qiu, B. Shen, M. Zhao, Z. Wang, B. Xie, and Y. Xu, “A nationwide survey of psychological distress among Chinese people in the COVID-19 epidemic: Implications and policy recommendations,” Gen. Psychiatry, vol. 33, no. 2, pp. 19–21, 2020.

[8] L. Cori, F. Bianchi, E. Cadum, and C. Anthonj, “Risk Perception and COVID-19,” Int. J. Environ. Res. Public Health, vol. 17, no. 9, p. 3114, Apr. 2020.

[9] P. M. Sandman, N. D. Weinstein, and W. K. Hallman, “Communications to Reduce Risk Underestimation and Overestimation,” Risk Decis. Policy, vol. 3, no. 2, pp. 93–108, 1998.

[10] R. Laxminarayan and A. Malani, “Economics of Infectious Diseases,” Oxford Handb. Heal. Econ., no. April, pp. 1–20, 2012.

[11] WHO, “Strategic preparedness and response plan,” 2020.

[12] Government of Nepal, “Corona Info-Ministry of Health and Population, Nepal,” 2020. [Online]. Available: https://covid19.mohp.gov.np/#/.

[13] R. R. Parajuli, “Citizen Disaster Science Education for effective disaster risk reduction in developing countries,” Geoenvironmental Disasters, vol. 7, no. 12, 2020.

[14] S. R. Baker, N. Bloom, S. J. Davis, and Stephen J. Terry, “COVID-INDUCED ECONOMIC UNCERTAINTY,” 2020.

[15] N. Fernandes, “Economic effects of coronavirus outbreak (COVID-19) on the world economy Nuno Fernandes Full Professor of Finance IESE Business School Spain,” 2020.

[16] UNDRR, “Leave no one hehind in COVID-19 prevention, response and recovery,” 2020.

[17] C. Hevia and A. Neumeyer, “A Conceptual Framework for Analyzing the Economic Impact of COVID-19 and its Policy Implications,” 2020.

[18] H. Inoue and Y. Todo, “The Propagation of the Economic Impact through Supply Chains: The Case of a Mega-City Lockdown against the Spread of COVID-19,” SSRN Electron. J., pp. 1–11, 2020.

[19] J. van der Pligt, “Risk Perception and Self-Protective Behavior,” Eur. Psychol., vol. 1, no. 1, pp. 34–43, 1996.

[20] R. A. Ferrer, W. M. P. Klein, A. Avishai, K. Jones, M. Villegas, and P. Sheeran, “When does risk perception predict protection motivation for health threats? A person-by-situation analysis,” PLoS One, vol. 13, no. 3, pp. e0191994–e0191994, Mar. 2018.

[21] W. J. Guan et al., “Clinical Characteristics of Coronavirus Disease 2019 in China,” N. Engl. J. Med., 2020.

[22] F. Zhou et al., “Clinical course and risk factors for mortality of adult inpatients with COVID-19 in Wuhan, China: a retrospective cohort study,” Lancet, vol. 395, no. 10229, pp. 1054–1062, 2020.

[23] S. Garg et al., “Hospitalization Rates and Characteristics of Patients Hospitalized with,” 2020.

[24] A. B. Docherty et al., “Features of 16,749 hospitalised UK patients with COVID-19 using the ISARIC WHO Clinical Characterisation Protocol,” medRxiv, 2020.

[25] NHS England, “COVID-19 Daily Deaths,” 2020. [Online]. Available: https://www.england.nhs.uk/statistics/statistical-work-areas/covid-19-daily-deaths/.

[26] R. Chatterjee, S. Bajwa, D. Dwivedi, R. Kanji, M. Ahammed, and R. Shaw, “COVID-19 Risk Assessment Tool: Dual application of risk communication and risk governance,” Prog. Disaster Sci., vol. 7, p. 100109, 2020.

[27] UNDP NEPAL, “Rapid Assessment of the Social and Economic Impacts of COVID-19 on the vulnerable groups in Nepal,” Kathmandu, 2020.

[28] G. Onder, G. Rezza, and S. Brusaferro, “Case-Fatality Rate and Characteristics of Patients Dying in Relation to COVID-19 in Italy,” JAMA - J. Am. Med. Assoc., 2020.

[29] Government of Spain, “Corona Virus disease (COVID-19) Update No: 92,” 2020.

[30] RKI, “Coronavirus Disease 2019 (COVID-19) Dialy Situation Report of the RKI (May 1st 2020),” 2020.

[31] NYC Health, “Coronavirus disease 2019 (COVID-19) daily data summary,” New York, 2020.

[32] S. Lakshmi Priyadarsini and M. Suresh, “Factors influencing the epidemiological characteristics of pandemic COVID 19: A TISM approach,” Int. J. Healthc. Manag., vol. 0, no. 0, pp. 1–10, 2020.

[33] P. Sharma, B. Guha-Khasnobis, and D. Raj Khanal, “Nepal human development report 2014: Beyond Geography, Unlocking Human Potential,” 2014.

[34] Central Bureao of Statistics, “National Population and Housing Census 2011 (National Report) Government of Nepal,” 2012.

[35] MOHP GoN, “Glimpses of department of health services annual report 2016/17,” Kathmandu, 2017.

[36] J. M. Jin et al., “Gender Differences in Patients With COVID-19: Focus on Severity and Mortality,” Front. Public Heal., vol. 8, no. April, pp. 1–6, 2020.

[37] T. Barbieri, G. Basso, and S. Scicchitano, “Italian Workers at Risk during the COVID-19 Epidemic,” SSRN Electron. J., vol. 21, no. 1, pp. 1–9, 2020.

[38] F. Y. Lan, C. F. Wei, Y. T. Hsu, D. C. Christiani, and S. N. Kales, “Work-related COVID-19 transmission in six Asian countries/areas: A follow-up study,” PLoS One, vol. 15, no. 5, pp. 1–11, 2020.

[39] D. K. Chu et al., “Physical distancing, face masks, and eye protection to prevent person-to-person transmission of SARS-CoV-2 and COVID-19: a systematic review and meta-analysis.,” Lancet, vol. 6736, no. 20, pp. 1–15, 2020.

[40] J. B. Dowd et al., “Demographic science aids in understanding the spread and fatality rates of COVID-19,” Proc. Natl. Acad. Sci., p. 202004911, 2020.

[41] H. A. Linstone and M. Turoff, The Delphi method: Techniques and Application. London, 1975.

[42] C. Okoli and S. D. Pawlowski, “The Delphi method as a research tool: an example, design considerations and applications,” Inf. Manag., vol. 42, no. 1, pp. 15–29, 2004.

[43] Central Bureau of Statistics, “Annual Household Survey 2015/2016,” Kathmandu, 2016.

[44] S. A. Lauer et al., “The Incubation Period of Coronavirus Disease 2019 (COVID-19) From Publicly Reported Confirmed Cases: Estimation and Application,” Ann. Intern. Med., vol. 172, no. 9, pp. 577–582, Mar. 2020.

[45] M. U. G. Kraemer et al., “Spread of yellow fever virus outbreak in Angola and the Democratic Republic of the Congo 2015–16: a modelling study,” Lancet Infect. Dis., vol. 17, no. 3, pp. 330–338, 2017.

